# Novel type 2 diabetes prediction score based on traditional risk factors and circulating metabolites: Model derivation and validation in two large cohort studies

**DOI:** 10.1101/2024.06.12.24308860

**Authors:** Ruijie Xie, Christian Herder, Sha Sha, Lei Peng, Hermann Brenner, Ben Schöttker

## Abstract

**Abstract:** 

**Importance:** The predictive value of metabolomics quantified by nuclear magnetic resonance (NMR) for type 2 diabetes risk has not been assessed before. In addition, previous studies with other metabolomics quantification methods did not have an external validation cohort, which leaves doubts about the robustness of the derived models in other settings.

**Objective:** This project aimed to evaluate the incremental predictive value of metabolomic biomarkers for assessing the 10-year risk of type 2 diabetes when added to the clinical Cambridge Diabetes Risk Score (CDRS), which includes HbA_1c_.

**Design, Setting, and Participants:** We utilized 60,362 participants of the UK Biobank (UKB) for model derivation, 25,870 participants of the UKB for internal validation and 4,383 participants from the German ESTHER cohort for external validation.

**Exposures:** A total of 249 NMR-derived metabolites were quantified using nuclear magnetic resonance (NMR) spectroscopy.

**Main Outcomes and Measures:** The main outcome was 10-year type 2 diabetes incidence.

**Results:** Eleven metabolomic biomarkers, including glycolysis-related metabolites, ketone bodies, amino acids, and lipids, were selected with LASSO regression. In internal validation within the UKB, adding these metabolites Harrel’s C-index of the clinical CDRS from 0.815 to 0.834 (*P*<0.001) and the continuous net reclassification index (NRI) was 39.8% (*P*<0.001). External validation in the ESTHER cohort showed a comparable C-index increase from 0.770 to 0.798 (*P*<0.001) and a continuous NRI of 33.8% (*P*<0.001).

**Conclusions and Relevance:** Adding 11 biomarkers, mainly from glucose and lipid metabolism, to the clinical CDRS led to a novel type 2 diabetes prediction model, the “UK Biobank Diabetes Risk Score” (UKB-DRS), which substantially outperformed the clinical CDRS. As only very limited clinical information and a blood sample are needed for the UKB-DRS, and as high-throughput NMR metabolomics are becoming increasingly available at low costs, this model has considerable potential for routine clinical application in diabetes risk assessment.

**Key Points:** *Question:* Can the inclusion of metabolites measured in blood samples with NMR spectroscopy improve the accuracy of the clinical Cambridge Diabetes Risk Score (CDRS), which is already a good prediction model including the main diabetes risk indicator HbA_1c_?

*Findings:* The novel UK Biobank Diabetes Risk Score (UKB-DRS), which includes traditional diabetes risk factors and 11 metabolites, demonstrated significantly enhanced predictive performance compared to the clinical CDRS, both in the UK Biobank and the German ESTHER cohort.

*Meaning:* The novel UKB-DRS could significantly improve the validity of early identification of individuals at risk for type 2 diabetes and guide clinicians and people at risk of diabetes in the choice of preventive measures.

## Introduction

The global prevalence of type 2 diabetes is on a significant upward trend, associated with increased mortality, diminished quality of life, and substantial economic burden.^1–3^ Early identification of individuals at elevated risk is essential, given the effectiveness of preventative measures in mitigating or delaying disease onset.^4^ Although current models for predicting type 2 diabetes risk effectively differentiate between individuals with low and high future risks, their clinical applicability is limited by a lack of specificity and an incomplete representation of the complex risk factors.^5–7^

Recent advances in metabolomics, particularly through the use of nuclear magnetic resonance (NMR) spectroscopy, offer promising insights into the early detection of type 2 diabetes.^8, 9^ The comprehensive metabolomic profiling enabled by NMR spectroscopy, including its ability to measure a wide array of metabolites in a single assay, provides a more nuanced view of the metabolic disturbances preceding type 2 diabetes.^10, 11^ Furthermore, the high-throughput nature of NMR spectroscopy, coupled with its low operational costs and minimal batch-to-batch variation, makes it an ideal tool for large-scale epidemiological studies.^12^

Despite the potential of metabolomics to enhance risk prediction models, previous studies have often been limited to investigating a small number or single subclasses of metabolomic biomarkers, leaving the value of metabolomic analysis in predicting the risk of type 2 diabetes uncertain.^13–16^ This study is the first aiming to derive and externally validate a NMR metabolomics data based risk score for type 2 diabetes in two large, population-based cohort studies.

## Methods

### Study population

The UK Biobank (UKB) is a large prospective cohort study with 502,493 participants, aged 37 to 73 years, recruited from 13 March 2006 to 1 October 2010 across 22 assessment sites in England, Scotland, and Wales.^17^

The ESTHER study is an ongoing population-based cohort study conducted in Saarland, a federal state in South-West Germany, with 9,940 participants, aged 50 to 75 years. The recruitment occurred during standard health checkups by general practitioners (GPs) from 1 July 2000 to 30 June 2002. Follow-ups were conducted every two to three years thereafter.^18^ The inclusion and exclusion criteria for the analyzed study population are shown in **Supplemental Text S1** and **Supplemental Figure S1.**

### Metabolomic profiling

Nightingale Health’s high-throughput NMR metabolomics platform was used to measure 250 metabolites from randomly selected baseline plasma samples of UKB participants, alongside all baseline serum samples from the ESTHER cohort with sufficient blood sample quality.^19^ Glycerol was excluded because it could not be measured in most of the participants of both cohorts, leaving n=249 metabolites for the analyses. The nomenclature and completeness of these metabolites are shown in **Supplemental Table S1**.

### Variables of the clinical CDRS and type 2 incidence ascertainment

The CDRS is a predictive tool used to assess the risk of individuals for future type 2 diabetes development. This scoring system includes age, sex, body mass index (BMI), family history of diabetes, smoking status, prescription of anti-hypertensive medication and steroids.^20^ If blood samples can be taken, the clinical CDRS is recommended to use, which additionally includes the HbA_1c_.^21^ The assessment methods of the variables of the clinical CDRS and of type 2 incidence in both cohorts are shown in **Supplemental Text S2** and **Supplemental Table S2**.

### Statistical analyses

#### General remarks

All analyses were performed using R software version 4.3.0 (R Foundation for Statistical Computing, Vienna, Austria), and statistical significance for two-sided tests was set at a *P*-value of <0.05. Missing values were imputed using the Random Forest estimation method in the r package *missForest* (version 1.5).^22^ The completeness of each variable within the UKB and ESTHER cohorts is detailed in **Supplemental Table S1**.

#### Metabolite selection and model derivation

First, concentrations of all metabolites were log-transformed to approximate a normal distribution for analysis. Subsequently, these values were independently scaled to standard deviation units within each cohort. The UKB cohort was divided into a derivation set (70%) and an internal validation set (30%). The ESTHER study served as the external validation cohort. The Least Absolute Shrinkage and Selection Operator (LASSO) is a regularization technique adept at identifying strong predictors among high-dimensional and correlated predictors, implemented through the ’*glmnet*’ package (version 4.1-7) in R.^23^ Ten-fold cross-validation, based on the lowest model validation error, was employed to determine the optimal tuning parameter λ for LASSO. A bootstrap LASSO method was then utilized, involving the creation of 1000 derivation sets and applying the LASSO procedure to each resampled dataset.^24^ Metabolites selected by LASSO in at least 95% of instances were designated as metabolites of interest, a threshold determined by a previous study to enhance model generalizability and minimize overfitting.^25^ These selected metabolites were subsequently incorporated into the clinical CDRS to construct a new prediction model.

#### Validation of the model’s predictive performance

The 30% UKB subsample was utilized as an internal validation cohort, and the ESTHER study served as an external validation cohort to evaluate the predictive performance of the derived models. The model’s predictive performance was assessed with discrimination, risk stratification and model calibration statistics. Discrimination was assessed using Harrell’s C-index and the area under the receiver operating characteristic curve (AUC). To determine whether the addition of metabolites improved model discrimination compared to the clinical CDRS, the statistical significance of improvements in the C-index was determined using the ’*compareC*’ package (version 1.3.2) in R. Additionally, the incremental discrimination ability of each metabolite was evaluated in the internal and validation set. The net reclassification index (NRI) and the integrated discrimination improvement index (IDI) were assessed to evaluate risk reclassification.^26^ Calibration of the predicted probabilities was performed using the Platt Scaling method, comparing the observed incidence rate of type 2 diabetes events in deciles of absolute predicted risk to their corresponding predicted event rates.^27^

#### Associations of selected metabolites with incident type 2 diabetes

To derive the hazard ratios (HRs) and 95% confidence intervals (CIs) of selected metabolites (per one standard deviation increment) for 10-year type 2 diabetes risk, metabolites were individually added to Cox proportional hazards regression models in the internal and the external validation cohort. These models were adjusted for the variables of the CDRS, using the r-package *survival* (version 3.5-5).

## Results

### Baseline characteristics

**Table 1** summarizes the characteristics of the clinical CDRS variables among 86,232 participants in the UKB cohort and 4,383 participants in the ESTHER study. The average age and sex distribution were similar across the two cohorts, with participants in the UKB having an average age of 59.9 years (SD 4.4) and comprising 44.3% males. In the ESTHER cohort, the mean age was 60.2 years (SD 5.5), with 42.7% of the participants being male. Furthermore, the BMI and HbA_1c_ levels were comparable. Compared to the UKB, a higher proportion of participants in the ESTHER study were current smokers (17.5% in ESTHER vs. 9.3% in the UKB), more had a family history of diabetes (36.9% vs. 16.8%) and more were prescribed anti-hypertensive medication (37.8% in ESTHER vs. 13.3% in the UKB). Steroid prescriptions were equally rare in both cohorts (0.8% in ESTHER vs. 1.3% in UKB).

**Table 1.** Baseline characteristics of selected participants from the UK Biobank and ESTHER study.

| Baseline characteristics | Derivation cohort | Validation cohort |
| --- | --- | --- |
|  | UK Biobank | ESTHER |
| Number of participants | 86,232 | 4,383 |
| Male sex, n (%) | 39,442 (44.3) | 1,871 (42.7) |
| Age (years), mean (SD) | 59.9 (5.4) | 60.2 (5.5) |
| HbA<sub>1c</sub> (%), mean (SD) | 3.1 (0.4) | 3.2 (0.5) |
| BMI (kg/m<sup>2</sup>), mean (SD) | 27.4 (4.5) | 27.4 (4.3) |
| BMI category, n (%) |  |  |
| <25 kg/m<sup>2</sup> | 27,626 (32.0) | 1,302 (29.7) |
| 25-27.49 kg/m<sup>2</sup> | 21,529 (25.0) | 1,193 (27.2) |
| 27.5-29.99 kg/m<sup>2</sup> | 16,666 (19.3) | 958 (21.7) |
| ≥ 30 kg/m<sup>2</sup> | 20,411 (23.7) | 984 (22.4) |
| Cigarette smoking, n (%) |  |  |
| Never | 46,150 (53.5) | 2,233 (50.9) |
| Ex-smoker | 32,075 (37.2) | 1,385 (31.6) |
| Current smoker | 8,007 (9.3) | 765 (17.5) |
| Family history of diabetes, n (%) |  |  |
| None | 71,783 (83.2) | 2,768 (63.2) |
| Parent or sibling | 12,982 (15.1) | 1,384 (31.6) |
| Parent and sibling | 1,467 (1.7) | 231 (5.3) |
| Prescribed medication, n (%) |  |  |
| Anti-hypertensive | 11,502 (13.3) | 1,658 (37.8) |
| Steroid | 1,104 (1.3) | 33 (0.8) |

**Abbreviations:** BMI, body mass index; HbA<sub>1c</sub>, glycated hemoglobin; SD, standard deviation.

### Associations of selected metabolites with incident type 2 diabetes

Over a follow-up of up to 10 years, 3,537 of 86,232 study participants of the UKB developed diabetes (incidence rate (IR) per 10,000 person-years (PY), 55.0) and 495 of 4,383 participants of the ESTHER study (IR per 10,000 PY, 145.2).

Through LASSO analyses and bootstrapping, 11 metabolites were selected to enhance the predictive power of the clinical CDRS for the risk of type 2 diabetes in the derivation set. These metabolites include four glycolysis related metabolites (citrate, glucose, lactate and pyruvate), two ketone bodies (3-hydroxybutyrate and acetate), two amino acids (glutamine and tyrosine), two lipoprotein related metabolites (IDL-CE-pct (cholesteryl esters to total lipids in IDL percentage) and M-LDL-TG-pct (triglycerides to total lipids in medium LDL percentage), and a fatty acid-related metabolite LA-pct (linoleic acid to total fatty acids percentage).

Figure 1 presents the associations between the 11 selected metabolites and incident type 2 diabetes within the internal validation set of the UKB and the ESTHER cohort, adjusted for the variables of the clinical CDRS graphically while the HRs with 95%CIs per SD increments and p-values can be found in **Supplemental Table S3**. All biomarkers except the ketone bodies 3-hydroxybutyrate and acetate were statistically significantly associated with type 2 diabetes in the UKB. Except for 3-hydroxyruvate, for which confidence intervals did not overlap, the results were remarkably similar in the ESTHER study. Results were also comparable for citrate and pyruvate although these biomarkers lacked statistical significance in the ESHTER study.

**Figure 1.**
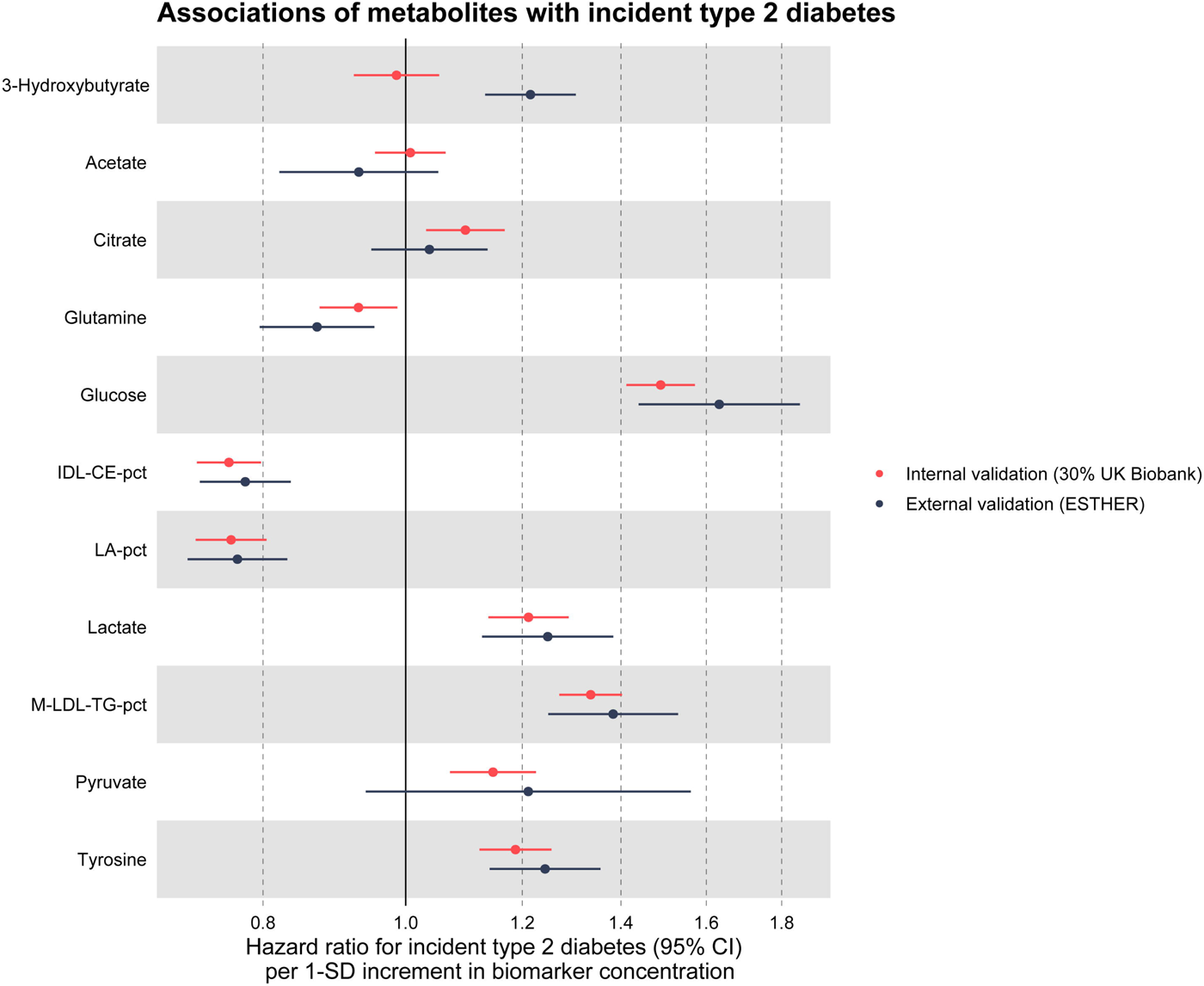
Associations between selected metabolites and incident type 2 diabetes in the internal validation cohort (30% of UK Biobank, N=25,870) and external validation cohort (ESTHER, N=5,904) Hazard ratios are expressed per 1 standard deviation of the respective metabolite concentration and are adjusted for age, sex, body mass index, smoking status, family history of diabetes, prescription of anti-hypertensive drugs, prescription of steroids and glycated hemoglobin. The exact HRs with 95%CIs and p-values per SD increments and the SDs of the metabolites are shown in Supplemental Table S3. Abbreviations: CI, confidence interval; IDL-CE-pct, cholesteryl esters to total lipids in IDL percentage; LA-pct, linoleic acid to total fatty acids percentage; M-LDL-TG-pct, triglycerides to total lipids in medium LDL percentage; SD, standard deviation.

### Improvements in type 2 diabetes risk prediction by metabolomic biomarkers

In the derivation set, the C-index for the clinical CDRS (0.830) was much higher than for the CDRS (0.740) (**Table 2**). Thus, we compared the predictive performance of the novel model of the clinical CDRS extended by 11 metabolites to the clinical CDRS, only. We call the novel model “UK Biobank Diabetes Risk Score (UKB-DRS)” in the following. Coefficients for all variables in the UKB-DRS are listed in **Supplemental Table S4**. In the derivation, internal validation and external validation set, the UKB-DRS consistently had a statistically significantly (*P*<0.001) higher C-index (95%CI) than the clinical CDRS, with comparable C-index increases of 0.017, 0.019 and 0.028 units, respectively. The receiver operating curves (ROC) are shown in **Supplemental Figure S2**. Moreover, an improved risk stratification ability was observed, with a continuous NRI of 39.8% (34.6%, 45.0%) and an IDI of 0.0216 (0.0170, 0.0262) in internal validation and a continuous NRI of 33.8% (26.4%, 41.2%), and an IDI of 0.0016 (0.0012, 0.0019) in external validation. The model calibration of both the clinical CDRS and the UKB-DRS was good in the internal and the external validation set and the confidence interval bands overlapped (**Supplemental Figure S3**).

**Table 2.** Comparison of the predictive performance of the clinical Cambridge Diabetes Risk Score and the novel developed UK Biobank Diabetes Risk Score for 10-year type 2 diabetes risk prediction.

| Metrics |  |
| --- | --- |
| **Derivation set (70% of UK Biobank)** |  |
| **Sample size / incident type 2 diabetes** | **N=60,362 / 2,477** |
| C-index (Clinical CDRS<sup>a</sup>) | 0.830 (0.821, 0.839) |
| C-index (UKB-DRS<sup>b</sup>); *P*-value<sup>c</sup> | 0.847 (0.838, 0.855); *P*<0.001 |
| **Internal validation set (30% of UK Biobank)** |  |
| **Sample size / incident type 2 diabetes** | **N=25,870 / 1,059** |
| C-index (Clinical CDRS<sup>a</sup>) | 0.815 (0.800, 0.829) |
| C-index (UKB-DRS<sup>b</sup>); *P*-value<sup>c</sup> | 0.834 (0.820, 0.847); *P*<0.001 |
| Continuous NRI (%); *P*-value<sup>c</sup> | 39.8 (34.6, 45.0); *P*<0.001 |
| IDI; *P*-value | 0.0216 (0.0170, 0.0262); *P*<0.001 |
| **External validation set (ESTHER)** |  |
| **Sample size / incident type 2 diabetes** | **N=4,383 / 495** |
| C-index (Clinical CDRS<sup>a</sup>) | 0.770 (0.750, 0.791) |
| C-index (UKB-DRS<sup>b</sup>); *P*-value<sup>c</sup> | 0.798 (0.779, 0.817); *P*<0.001 |
| Continuous NRI (%) (UKB-DRS<sup>b</sup>); *P*-value<sup>c</sup> | 33.8 (26.4, 41.2); *P*<0.001 |
| IDI (UKB-DRS<sup>b</sup>); *P*-value<sup>c</sup> | 0.0016 (0.0012, 0.0019); *P*<0.001 |

**Abbreviations:** CDRS, Cambridge Diabetes Risk Score; IDI, integrated discrimination improvement; NRI, net reclassification improvement, UKB-DRS (UK Biobank Diabetes Risk Score)
<sup>a</sup> The clinical Cambridge Diabetes Risk Score (CDRS) consists of the following diabetes risk factors: (age, sex, BMI, smoking status, family history of diabetes, prescription of anti-hypertensive medication and steroids, and HbA<sub>1c</sub>).
<sup>b</sup> The UK Biobank Diabetes Risk Score (UKB-DRS) is based on the traditional diabetes risk factors of the clinical CDRS (see above) and the following 11 metabolites: glucose, citrate, lactate, pyruvate, 3-hydroxybutyrate, acetate, glutamine, tyrosine, IDL-CE-pct, LA-pct, and M-LDL-TG-pct.
<sup>c</sup> *P*-value for comparison with the clinical CDRS

Figure 2 illustrates the incremental improvement in the C-index for each of the 11 selected metabolites after their incorporation into the clinical CDRS in internal and external validation. Especially glucose, IDL-CE-pct, LA-pct and M-LDL-TG-pct enhanced the discriminative ability of the model in both cohorts. The other seven metabolites showed low increases in C-statistic at best and their results were not always consistent in the two cohorts. Thus, as a post hoc analysis, we also assessed the C-index of the clinical CDRS extended by 4 metabolites only (glucose, IDL-CE-pct, LA-pct and M-LDL-TG-pct with the coefficients shown in **Table S4**). With C-index (95%CI) values of 0.830 (0.815, 0.844) and 0.786 (0.766, 0.805) in the internal and external validation set, respectively, the predictive performance was almost as good as the one of the model including 11 metabolites in the internal validation (C-index: 0.834) and a little weaker in the external validation (C-index: 0.798).

**Figure 2.**
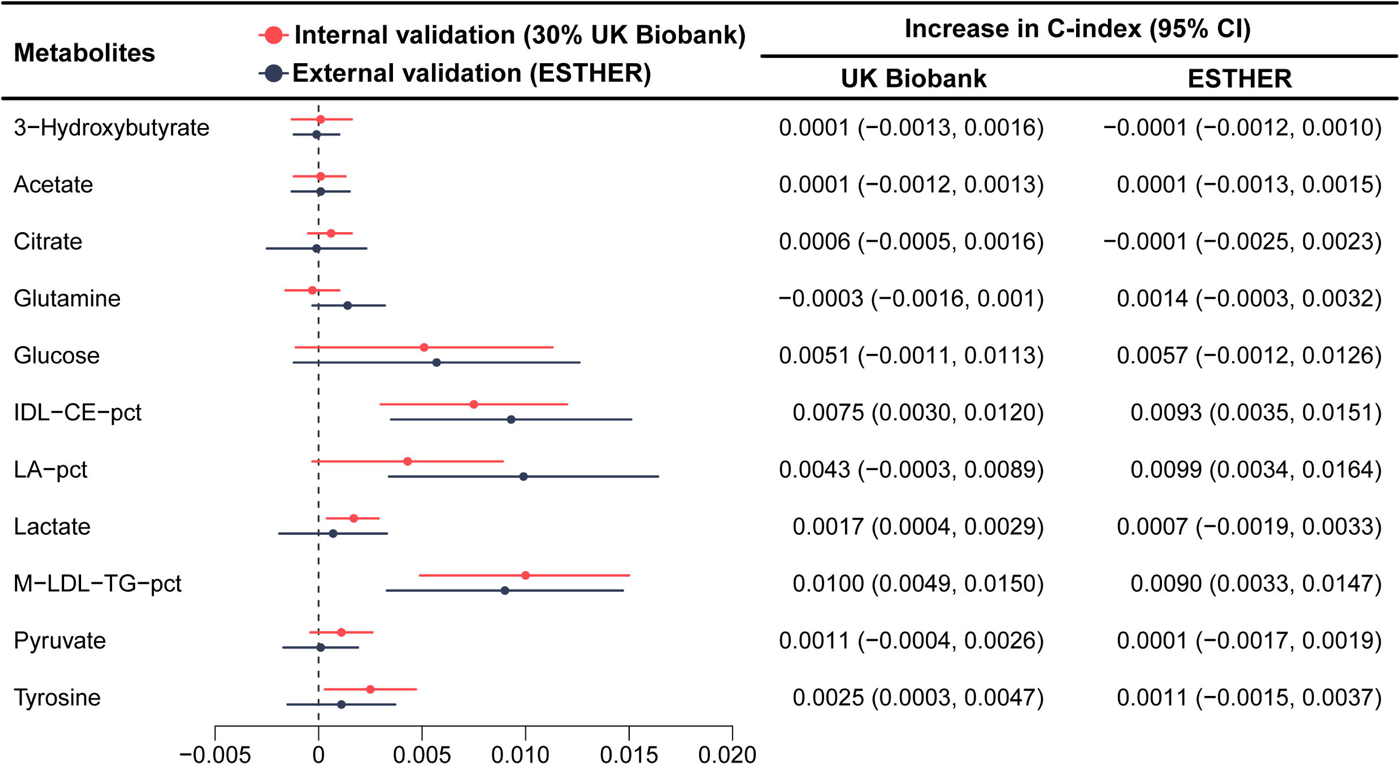
The incremental discrimination of each metabolite for the model after the selected metabolites were added to the clinical Cambridge Diabetes Risk Score in the internal validation cohort (30% of UK Biobank, N=25,870) Abbreviations: CI, confidence interval; IDL-CE-pct, cholesteryl esters to total lipids in IDL percentage; LA-pct, linoleic acid to total fatty acids percentage; M-LDL-TG-pct, triglycerides to total lipids in medium LDL percentage.

## Discussion

To our knowledge, this study represents the largest-scale investigation to date on the predictive utility of metabolomic biomarkers for type 2 diabetes risk, leveraging data from two large European cohorts comprising over 90,000 middle-aged and older adults of whom more than 4,000 developed type 2 diabetes during 10 years of follow-up. We developed and validated the UKB-DRS that integrates an established risk prediction model (clinical CDRS) with 11 metabolites for enhanced predictive performance of 10-year type 2 diabetes risk. In both internal and external validation, the UKB-DRS demonstrated significant improvements in the C-index compared to the clinical CDRS, with increases of 0.019 and 0.028, respectively. Additionally, we examined the incremental value of each metabolite to the model’s predictive capability across both cohorts, finding similar results in internal and external validation for all 11 metabolites. These findings confirm the robustness and generalizability of the new algorithm in enhancing the predictive accuracy for the risk of type 2 diabetes.

### Previous studies & Interpretation of findings

To enhance the accuracy of existing prediction models for type 2 diabetes risk, numerous studies have investigated the incremental value of metabolomic biomarkers.^28^ Many traditional prediction models already achieve high predictive accuracy, making significant improvements upon these models challenging.^5, 6^ Previous studies often focused on the impact of specific types of metabolomic biomarkers, with several studies finding that incorporating pre-selected metabolomic biomarkers into traditional risk prediction models did not enhance risk identification capabilities.^29, 30^ However, other studies have shown that screening among a large number of metabolomic biomarkers tends to improve model discrimination.^15, 31^ The European Prospective Investigation into Cancer and Nutrition (EPIC)-Potsdam study, which measured 163 metabolites for 800 incident type 2 diabetes cases (with an average follow-up of 7 years) and 2,282 controls, showed that adding 14 metabolomic biomarkers selected from 163 metabolite candidates to a traditional risk model significantly improved the model’s discriminative ability (C-index increased from 0.901 to 0.912).^15^ The EPIC-Potsdam study is the second largest study after the UK Biobank using untargeted metabolomics data. Prior to our study, a study from the UKB, which did not have the full number of metabolomics data available had the largest sample size (n=65,684 participants analyzed, including 1,719 type 2 diabetes cases).^31^ The authors conducted internal validation after incorporating metabolomic data into the Framingham Diabetes Risk Score, significantly enhancing the model’s C-index from 0.802 to 0.830 (Δ C-index=0.028, *P*<0.001). However, a lack of external validation remains a major barrier to translating metabolomic based models into clinical applications, which we overcame by our joint analysis of the UKB and ESTHER cohort. Furthermore, we included more than twice the number of incident type 2 diabetes case in our UKB analysis (n=3,536) compared to the previous UKB study by including more study participants with metabolomics data (n=86,232) and by the additional use of primary care records for the outcome assessment.

### Biological mechanisms of selected metabolites

The ensemble of metabolites identified in our study captures a broad spectrum of biochemical pathways integral to the metabolic dysregulation that precedes type 2 diabetes. The identified metabolites stem from the tricarboxylic acid cycle (citrate, glucose, lactate and pyruvate), are ketone bodies (3-hydroxybutyrate and acetate), amino acids (glutamine and tyrosine), lipoproteins (IDL-CE-pct and M-LDL-TG-pct) and a fatty acid (LA-pct). This metabolic signature characterizes an impaired glucose and lipid metabolism that is detectable in blood samples up to 10 years prior diabetes diagnosis and might also give new insights into the mechanisms of type 2 diabetes pathogenesis.

Citrate is a key metabolite in the tricarboxylic acid cycle and not only facilitates energy production but also plays a vital role in fatty acid synthesis, thereby linking glycolytic processes with lipid metabolism.^9^ Concurrently, glucose as a direct participant in glycolysis, provides critical insights into the disrupted glucose homeostasis that is characteristic of diabetes.^32^ The presence of lactate and pyruvate, products of anaerobic and aerobic glycolysis respectively, signals a shift towards inefficient energy utilization, a common observation in insulin-resistant states. These shifts are believed to reflect the underlying mitochondrial adaptations that compromise cellular energy management and exacerbate hyperglycemic conditions.^33^

Ketone bodies, including 3-hydroxybutyrate and acetate, accumulate during increased fatty acid oxidation - a compensatory response to impaired glucose utilization in diabetes. Elevated levels of these ketone bodies indicate the body’s attempt to maintain energy balance under conditions of insulin resistance.^34^

Furthermore, the amino acids glutamine and tyrosine are noted for their dual roles in metabolic and neurotransmitter pathways. Glutamine is involved in gluconeogenesis and plays a crucial role in maintaining glucose homeostasis. Tyrosine is associated with catecholamine synthesis and may influence stress responses that exacerbate beta-cell dysfunction and insulin resistance.^32^

Lipoprotein-related metabolites, including IDL-CE-pct and M-LDL-TG-pct, reflect changes in lipoprotein metabolism that contribute to a dyslipidemia pattern commonly associated with insulin resistance.^35^ Additionally, the percentage of linoleic acid, a crucial component in inflammatory pathways, has been associated with variations in dietary intake and metabolic status, influencing cellular functions pivotal to insulin sensitivity.^36^

Overall, the identified metabolic profile comprising glycolysis-related metabolites, ketone bodies, amino acids, lipoprotein-related metabolites and fatty acids not only contributes evidence to the biological pathways from early metabolic disturbances to type 2 diabetes development but also adds predictive power to traditional risk factor models, which will allow to detect those better who are at risk to develop type 2 diabetes and profit most from early preventive interventions.

### Strengths and limitations

This study stands out from previous research primarily due to its substantial sample size and the confirmation of our model’s robustness through external validation. We employed a well-established, clinically approved targeted NMR metabolomics platform with absolute quantification of 250 biomarkers. This feature does not only facilitate comparative analyses across different populations but also enhances the potential for clinical translation.

However, our study is not without limitations. Although we only included UKB participants with available primary care records to ascertain the incidence of type 2 diabetes, the IR remained significantly lower than in the ESTHER study (55.0 vs 145.2 per 10,000 person-years, respectively). This difference is likely due to an underreporting in the UKB and a more complete ascertainment in the ESTHER study, in which the GPs of the study participants were asked to provide their medical records and these were screened for diabetes diagnoses and glucose-lowering drugs. In addition, there are differences between the two cohorts regarding the blood samples. The UKB used plasma and ESTHER serum samples to determine the metabolites. Furthermore, only a small proportion of blood samples in the UK Biobank cohort were collected under fasting conditions, defined as no consumption of food or drink for at least 8 hours (3.3%), whereas the majority of participants in the ESTHER study were not fasting (90.6%). However, a comparison of the levels of the 11 selected metabolites by fasting status in the UKB and ESTHER study shows that the differences in metabolite concentrations between fasting and non-fasting individuals are small in both cohorts (see **Supplemental Table S5**). This is in agreement with a previous study, which has shown that the duration of fasting has little impact on the variability of these metabolites.^37^ Thus, the novel UKB-DRS is robust towards fasting status and use of either serum or plasma samples. However, it is unknown how the model will perform in different populations. As its derivation and validation are based on British and German populations aged 50 to 69 years, extending its application to ethnically diverse populations or other age groups requires further validation.

## Conclusions

In conclusion, this study provides large-scale evidence from the UKB that a specific metabolomic profile indicative of alterations in glucose and lipid metabolism has additional value for the prediction of type 2 diabetes compared to a traditional risk factor-based model including the HbA_1c_. The derived novel UKB-DRS, which combines these traditional risk factors with 11 metabolomic biomarkers was robust in external validation using an independent, population-based German cohort. These findings have important future translational implications. With increasing clinical accessibility to high-throughput, targeted metabolomics analyses with the NMR technology, the use of these biomarkers in clinical risk prediction models is feasible. This can be useful for risk communication aiming at lifestyle changes and prioritizing cost-intensive preventive interventions (e.g., semaglutide injections for weight loss) to those at high risk for future type 2 diabetes development.

## Data Availability

All data produced in the present study are available upon reasonable request to the authors

## List of abbreviations

AUC: area under the receiver operating characteristic curve
CDRS: Cambridge Diabetes Risk Score
CI: confidence intervals
EPIC: European Prospective Investigation into Cancer and Nutrition
ESTHER: Epidemiologische Studie zu Chancen der Verhütung, Früherkennung und optimierten Therapie chronischer Erkrankungen in der älteren Bevölkerung
GlycA: glycoprotein acetyls
GP: general practitioner
HR: hazard ratio
IDI: integrated discrimination improvement
IDL-CE-pct: cholesteryl esters to total lipids in IDL percentage
IR: incidence rate
LA-pct: linoleic acid to total fatty acids percentage
LASSO: least absolute shrinkage and selection operator
M-LDL-TG-pct: triglycerides to total lipids in medium LDL percentage
NMR: nuclear magnetic resonance
NRI: net reclassification index
PY: person-years
SD: standard deviation
UKB: UK Biobank
UKB-DRS: UK Biobank-Diabetes Risk Score
VLDL-size: the average diameter for very low-density lipoprotein particles

## Acknowledgements

We would like to thank all participants of the ESTHER and UK Biobank cohort as well as the GPs of the ESTHER study and the staff of the UK Biobank assessment centers for their contribution to the studies this research is based on. Part of this research was conducted using the UK Biobank Resource under application 101633.

## Article Information

### Funding

The ESTHER study was funded by grants from the Baden-Württemberg state Ministry of Science, Research and Arts (Stuttgart, Germany), the Federal Ministry of Education and Research (Berlin, Germany), the Federal Ministry of Family Affairs, Senior Citizens, Women and Youth (Berlin, Germany), and the Saarland state ministry for Social Affairs, Health, Women and Family Affairs (Saarbrücken, Germany). UK Biobank was established by the Wellcome Trust, Medical Research Council, Department of Health, Scottish government, and Northwest Regional Development Agency. It has also had funding from the Welsh assembly government and the British Heart Foundation. The German Diabetes Center is funded by the German Federal Ministry of Health (Berlin, Germany) and the Ministry of Culture and Science of the state North Rhine-Westphalia (Düsseldorf, Germany) and receives additional funding from the German Federal Ministry of Education and Research (BMBF) through the German Center for Diabetes Research (DZD e.V.). The sponsors had no role in data acquisition or the decision to publish the data.

### Duality of Interest

The authors declare that they have no competing interests.

### Author’s Contributions

H.B. designed and led the ESTHER cohort. R.X. and B.S. generated the idea for the study and formulated the analytical plan. R.X. performed the data analyses and drafted the manuscript.

B.S. revised it. All authors contributed valuable intellectual content to the discussion. The corresponding author attests that all listed authors meet authorship criteria and that no others meeting the criteria have been omitted. R.X. and B.S. had full access to UK Biobank and ESTHER data used for this study and are the guarantors of the manuscript and accepts full responsibility for the work and/or the conduct of the study.

### Prior Presentation

Not applicable.

---

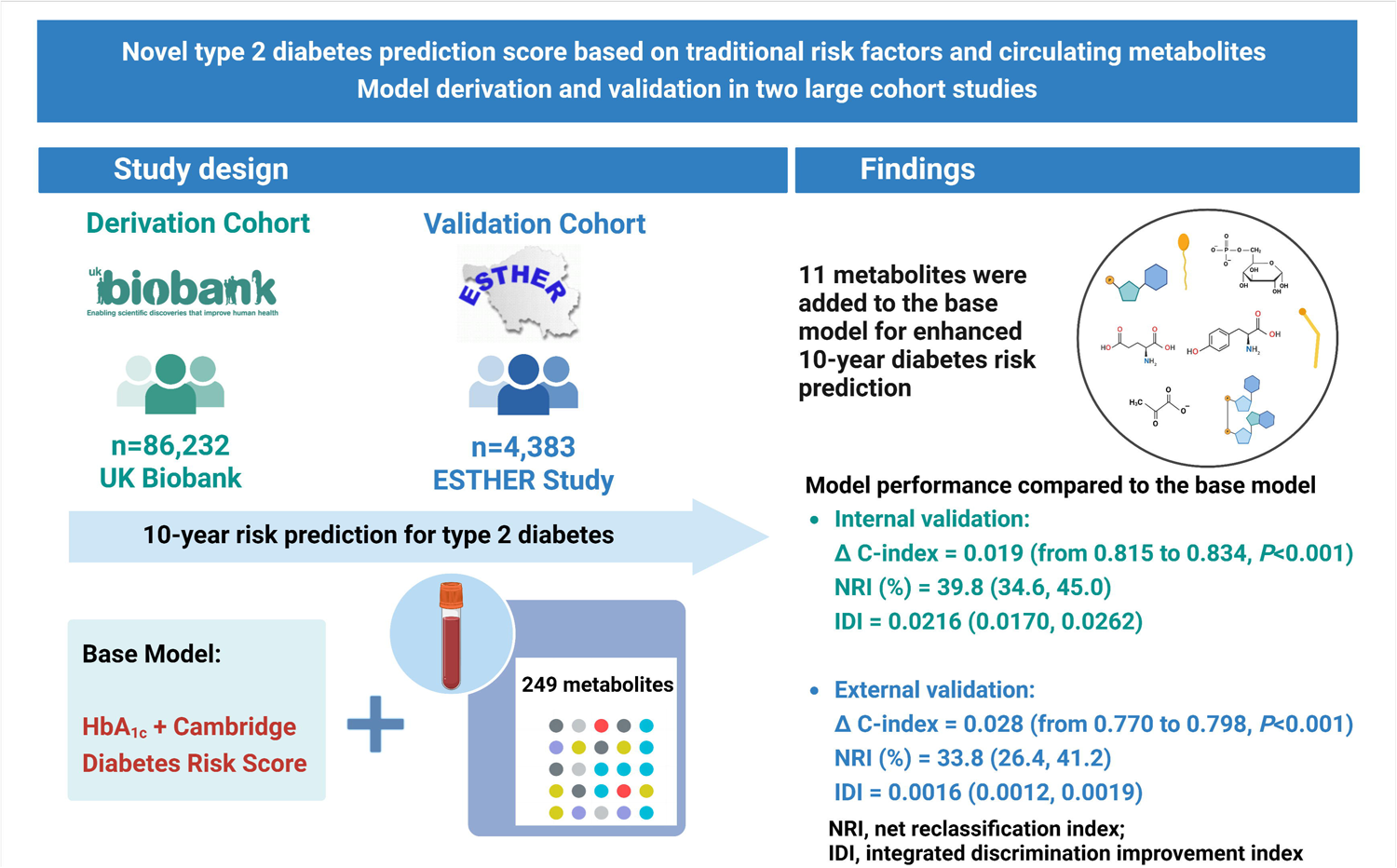

## Notes

### Competing Interest Statement

The authors have declared no competing interest.

### Funding Statement

The ESTHER study was funded by grants from the Baden-Wurttemberg state Ministry of Science, Research and Arts (Stuttgart, Germany), the Federal Ministry of Education and Research (Berlin, Germany), the Federal Ministry of Family Affairs, Senior Citizens, Women and Youth (Berlin, Germany), and the Saarland state ministry for Social Affairs, Health, Women and Family Affairs (Saarbrucken, Germany). UK Biobank was established by the Wellcome Trust, Medical Research Council, Department of Health, Scottish government, and Northwest Regional Development Agency. It has also had funding from the Welsh assembly government and the British Heart Foundation. The German Diabetes Center is funded by the German Federal Ministry of Health (Berlin, Germany) and the Ministry of Culture and Science of the state North Rhine-Westphalia (Dusseldorf, Germany) and receives additional funding from the German Federal Ministry of Education and Research (BMBF) through the German Center for Diabetes Research (DZD e.V.). The sponsors had no role in data acquisition or the decision to publish the data.

### Author Declarations

The UKB received ethical approval from the North-West Multicentre Research Ethics Committee (REC reference: 11/NW/03820). The ESTHER study was approved by the Ethics Committee of the Medical Faculty of the University of Heidelberg (Application number: S-58/2000). Both UKB and ESTHER study are conducted in accordance with the 1964 Helsinki declaration and its later amendments. All study participants of UKB and ESTHER study gave written informed consent.

