## Supplemental Materials for "Novel type 2 diabetes prediction score based on traditional risk factors and circulating metabolites: Model derivation and validation in two large cohort studies"

**Supplemental Text S1.** The inclusion and exclusion criteria of the analyzed study population from the UK Biobank and ESTHER study

In the UK Biobank (UKB) and ESTHER cohort n=274,353 and n=8,308 study participants were randomly selected for metabolomics measurements, respectively. Primary care records (encompassing GP clinical incident records, GP prescribing records and GP registration records) were only available for approximately 45% of the UKB participants. To minimize the potential underestimation of the incidence of type 2 diabetes within the UKB cohort, we excluded participants living in UK locations not covered by linkage with primary healthcare records of the NHS (see **Supplemental Figure S1** for study flow-charts). In the ESTHER study, participants whose blood samples were flagged with quality control warnings for low glucose or high pyruvate were also excluded. In the UKB, these quality control warnings related to time of storage of blood samples at room temperature were negligible. To ensure comparability in age distribution between the cohorts, participants not in the age range of 50-69 years were further excluded. Lastly, we excluded individuals with physician-diagnosed or self-reported diabetes diagnoses prior to the baseline of the cohorts, with potentially undiagnosed diabetes (HbA_1c_ ≥ 6.5% [48 mmol/mol] or taking glucose-lowering drugs) or with missing data on diabetes from both cohorts. After these exclusions, this study included n=86,232 participants without diabetes mellitus from the UKB and n=4,383 from the ESTHER study.

**Supplemental Text S2.** Assessment methods of the variables of the clinical Cambridge Diabetes Risk Score and the outcomes in the UK Biobank and ESTHER study

In both the UKB and ESTHER study, information on demographic characteristics, lifestyle factors and medical history, including age, sex, family history of diabetes and smoking status were collected by standardized questionnaires. In the UKB, prescribed medication was obtained from self-report, primary care records and verbal interviews. In the ESTHER study, GPs completed a standardized health check-up form and documented current drug prescriptions. Weight and height were measured in both studies – in the UKB by study staff in in the assessment centers and in the ESTHER study by GPs during the health check-up. The HbA_1c_ was measured from whole-blood samples with high-performance liquid chromatography on the Variant II (Bio-Rad) in both cohorts.

In the UKB, incident type 2 diabetes was ascertained by three sources.^1^ First, participants self-reported diagnoses of type 2 diabetes or use of glucose-lowering medications during follow-up surveys. Second, medical codes for type 2 diabetes in primary care records, hospital admissions, and death registry data were utilized for case identification. Prescription data for glucose-lowering medications from primary care were also used to identify cases (see **Supplemental Table S2** for diagnosis and medication codes).

In the ESTHER study, the ascertainment of incident type 2 diabetes was identified via four sources as described previously.^2^ Participants provided self-reports of diabetes diagnoses and glucose-lowering medication use via standardized questionnaires during the 2-, 5-, 8-, and 11-year follow-ups. Self-reported diagnoses at the 2- and 5-year marks were validated through questionnaires sent to the GPs of the participants, with unconfirmed cases being excluded. Similarly, at the 8- and 11-year follow-ups, GPs received questionnaires inquiring about new diabetes diagnoses within the last three years and unconfirmed self-reports were disregarded.

Study participants were censored at the first occurrence of a diabetes diagnosis, date of death, data of loss to follow-up, or when the maximum follow-up time of 10 years was reached.

**References to Supplemental Text S2**

1.	Bragg F, Trichia E, Aguilar-Ramirez D, Bešević J, Lewington S, Emberson J. Predictive value of circulating NMR metabolic biomarkers for type 2 diabetes risk in the UK Biobank study. *BMC Med*. May 3 2022;20(1):159. doi:10.1186/s12916-022-02354-9

2.	Schöttker B, Xuan Y, Gào X, Anusruti A, Brenner H. Oxidatively Damaged DNA/RNA and 8-Isoprostane Levels Are Associated With the Development of Type 2 Diabetes at Older Age: Results From a Large Cohort Study. *Diabetes Care*. Jan 2020;43(1):130-136. doi:10.2337/dc19-1379

**Supplemental** **Table S1.** Completeness of variables among the included study population from the UK Biobank and the ESTHER study before multiple imputation

| **Variable** | **UK Biobank  n (%)** | **ESTHER  n (%)** |
| --- | --- | --- |
| Age | 86232 (100.0%) | 4383 (100.0%) |
| Sex | 86232 (100.0%) | 4383 (100.0%) |
| BMI | 85933 (99.7%) | 4383 (100.0%) |
| Smoking status | 85929 (99.6%) | 4278 (97.6%) |
| Anti-hypertensive medication | 86232 (100.0%) | 4370 (99.7%) |
| Prescription of steroid | 86232 (100.0%) | 4383 (100.0%) |
| Family history of diabetes | 81028 (94.0%) | 4216 (96.2%) |
| HbA_1c_ | 82249 (95.4%) | 4343 (99.1%) |
| Acetate | 86168 (99.9%) | 4383 (100.0%) |
| Acetoacetate | 86228 (100.0%) | 4382 (100.0%) |
| Acetone | 86230 (100.0%) | 4383 (100.0%) |
| Ala | 86194 (100.0%) | 4355 (99.4%) |
| Albumin | 86223 (100.0%) | 4383 (100.0%) |
| ApoA1 | 86232 (100.0%) | 4383 (100.0%) |
| ApoB | 86232 (100.0%) | 4383 (100.0%) |
| ApoB-by-ApoA1 | 86232 (100.0%) | 4383 (100.0%) |
| Cholines | 86155 (99.9%) | 4379 (99.9%) |
| Citrate | 86224 (100.0%) | 4328 (98.7%) |
| Clinical-LDL-C | 86232 (100.0%) | 4383 (100.0%) |
| Creatinine | 83969 (97.4%) | 4050 (92.4%) |
| DHA | 86155 (99.9%) | 4379 (99.9%) |
| DHA-pct | 86155 (99.9%) | 4379 (99.9%) |
| Gln | 85992 (99.7%) | 4300 (98.1%) |
| Glucose | 86077 (99.8%) | 4378 (99.9%) |
| Gly | 86119 (99.9%) | 4356 (99.4%) |
| GlycA | 86231 (100.0%) | 4383 (100.0%) |
| HDL-C | 86232 (100.0%) | 4383 (100.0%) |
| HDL-CE | 86232 (100.0%) | 4383 (100.0%) |
| HDL-FC | 86232 (100.0%) | 4383 (100.0%) |
| HDL-L | 86232 (100.0%) | 4383 (100.0%) |
| HDL-P | 86232 (100.0%) | 4383 (100.0%) |
| HDL-PL | 86232 (100.0%) | 4383 (100.0%) |
| HDL-TG | 86232 (100.0%) | 4383 (100.0%) |
| HDL-size | 86232 (100.0%) | 4383 (100.0%) |
| His | 86111 (99.9%) | 4357 (99.4%) |
| IDL-C | 86232 (100.0%) | 4383 (100.0%) |
| IDL-CE | 86232 (100.0%) | 4383 (100.0%) |
| IDL-CE-pct | 86232 (100.0%) | 4383 (100.0%) |
| IDL-C-pct | 86232 (100.0%) | 4383 (100.0%) |
| IDL-FC | 86232 (100.0%) | 4383 (100.0%) |
| IDL-FC-pct | 86232 (100.0%) | 4383 (100.0%) |
| IDL-L | 86232 (100.0%) | 4383 (100.0%) |
| IDL-P | 86232 (100.0%) | 4383 (100.0%) |
| IDL-PL | 86232 (100.0%) | 4383 (100.0%) |
| IDL-PL-pct | 86232 (100.0%) | 4383 (100.0%) |
| IDL-TG | 86232 (100.0%) | 4383 (100.0%) |
| IDL-TG-pct | 86232 (100.0%) | 4383 (100.0%) |
| Ile | 86216 (100.0%) | 4383 (100.0%) |
| LA | 86155 (99.9%) | 4379 (99.9%) |
| LA-pct | 86155 (99.9%) | 4379 (99.9%) |
| LDL-C | 86232 (100.0%) | 4383 (100.0%) |
| LDL-CE | 86232 (100.0%) | 4383 (100.0%) |
| LDL-FC | 86232 (100.0%) | 4383 (100.0%) |
| LDL-L | 86232 (100.0%) | 4383 (100.0%) |
| LDL-P | 86232 (100.0%) | 4383 (100.0%) |
| LDL-PL | 86232 (100.0%) | 4383 (100.0%) |
| LDL-TG | 86232 (100.0%) | 4383 (100.0%) |
| LDL-size | 86232 (100.0%) | 4383 (100.0%) |
| L-HDL-C | 86232 (100.0%) | 4383 (100.0%) |
| L-HDL-CE | 86232 (100.0%) | 4383 (100.0%) |
| L-HDL-CE-pct | 86232 (100.0%) | 4383 (100.0%) |
| L-HDL-C-pct | 86232 (100.0%) | 4383 (100.0%) |
| L-HDL-FC | 86232 (100.0%) | 4383 (100.0%) |
| L-HDL-FC-pct | 86232 (100.0%) | 4383 (100.0%) |
| L-HDL-L | 86232 (100.0%) | 4383 (100.0%) |
| L-HDL-P | 86232 (100.0%) | 4383 (100.0%) |
| L-HDL-PL | 86232 (100.0%) | 4383 (100.0%) |
| L-HDL-PL-pct | 86232 (100.0%) | 4383 (100.0%) |
| L-HDL-TG | 86232 (100.0%) | 4383 (100.0%) |
| L-HDL-TG-pct | 86232 (100.0%) | 4383 (100.0%) |
| L-LDL-C | 86232 (100.0%) | 4383 (100.0%) |
| L-LDL-CE | 86232 (100.0%) | 4383 (100.0%) |
| L-LDL-CE-pct | 86232 (100.0%) | 4383 (100.0%) |
| L-LDL-C-pct | 86232 (100.0%) | 4383 (100.0%) |
| L-LDL-FC | 86232 (100.0%) | 4383 (100.0%) |
| L-LDL-FC-pct | 86232 (100.0%) | 4383 (100.0%) |
| L-LDL-L | 86232 (100.0%) | 4383 (100.0%) |
| L-LDL-P | 86232 (100.0%) | 4383 (100.0%) |
| L-LDL-PL | 86232 (100.0%) | 4383 (100.0%) |
| L-LDL-PL-pct | 86232 (100.0%) | 4383 (100.0%) |
| L-LDL-TG | 86232 (100.0%) | 4383 (100.0%) |
| L-LDL-TG-pct | 86232 (100.0%) | 4383 (100.0%) |
| L-VLDL-C | 86232 (100.0%) | 4383 (100.0%) |
| L-VLDL-CE | 86232 (100.0%) | 4383 (100.0%) |
| L-VLDL-CE-pct | 86232 (100.0%) | 4383 (100.0%) |
| L-VLDL-C-pct | 86232 (100.0%) | 4383 (100.0%) |
| L-VLDL-FC | 86232 (100.0%) | 4383 (100.0%) |
| L-VLDL-FC-pct | 86232 (100.0%) | 4383 (100.0%) |
| L-VLDL-L | 86232 (100.0%) | 4383 (100.0%) |
| L-VLDL-P | 86232 (100.0%) | 4383 (100.0%) |
| L-VLDL-PL | 86232 (100.0%) | 4383 (100.0%) |
| L-VLDL-PL-pct | 86232 (100.0%) | 4383 (100.0%) |
| L-VLDL-TG | 86232 (100.0%) | 4383 (100.0%) |
| L-VLDL-TG-pct | 86232 (100.0%) | 4383 (100.0%) |
| Lactate | 86062 (99.8%) | 4382 (100.0%) |
| Leu | 86224 (100.0%) | 4383 (100.0%) |
| MUFA | 86155 (99.9%) | 4379 (99.9%) |
| MUFA-pct | 86155 (99.9%) | 4379 (99.9%) |
| M-HDL-C | 86232 (100.0%) | 4383 (100.0%) |
| M-HDL-CE | 86232 (100.0%) | 4383 (100.0%) |
| M-HDL-CE-pct | 86232 (100.0%) | 4383 (100.0%) |
| M-HDL-C-pct | 86232 (100.0%) | 4383 (100.0%) |
| M-HDL-FC | 86232 (100.0%) | 4383 (100.0%) |
| M-HDL-FC-pct | 86232 (100.0%) | 4383 (100.0%) |
| M-HDL-L | 86232 (100.0%) | 4383 (100.0%) |
| M-HDL-P | 86232 (100.0%) | 4383 (100.0%) |
| M-HDL-PL | 86232 (100.0%) | 4383 (100.0%) |
| M-HDL-PL-pct | 86232 (100.0%) | 4383 (100.0%) |
| M-HDL-TG | 86232 (100.0%) | 4383 (100.0%) |
| M-HDL-TG-pct | 86232 (100.0%) | 4383 (100.0%) |
| M-LDL-C | 86232 (100.0%) | 4383 (100.0%) |
| M-LDL-CE | 86232 (100.0%) | 4383 (100.0%) |
| M-LDL-CE-pct | 86232 (100.0%) | 4383 (100.0%) |
| M-LDL-C-pct | 86232 (100.0%) | 4383 (100.0%) |
| M-LDL-FC | 86232 (100.0%) | 4383 (100.0%) |
| M-LDL-FC-pct | 86232 (100.0%) | 4383 (100.0%) |
| M-LDL-L | 86232 (100.0%) | 4383 (100.0%) |
| M-LDL-P | 86232 (100.0%) | 4383 (100.0%) |
| M-LDL-PL | 86232 (100.0%) | 4383 (100.0%) |
| M-LDL-PL-pct | 86232 (100.0%) | 4383 (100.0%) |
| M-LDL-TG | 86232 (100.0%) | 4383 (100.0%) |
| M-LDL-TG-pct | 86232 (100.0%) | 4383 (100.0%) |
| M-VLDL-C | 86232 (100.0%) | 4383 (100.0%) |
| M-VLDL-CE | 86232 (100.0%) | 4383 (100.0%) |
| M-VLDL-CE-pct | 86232 (100.0%) | 4383 (100.0%) |
| M-VLDL-C-pct | 86232 (100.0%) | 4383 (100.0%) |
| M-VLDL-FC | 86232 (100.0%) | 4383 (100.0%) |
| M-VLDL-FC-pct | 86232 (100.0%) | 4383 (100.0%) |
| M-VLDL-L | 86232 (100.0%) | 4383 (100.0%) |
| M-VLDL-P | 86232 (100.0%) | 4383 (100.0%) |
| M-VLDL-PL | 86232 (100.0%) | 4383 (100.0%) |
| M-VLDL-PL-pct | 86232 (100.0%) | 4383 (100.0%) |
| M-VLDL-TG | 86232 (100.0%) | 4383 (100.0%) |
| M-VLDL-TG-pct | 86232 (100.0%) | 4383 (100.0%) |
| Omega-3 | 86155 (99.9%) | 4379 (99.9%) |
| Omega-3-pct | 86155 (99.9%) | 4379 (99.9%) |
| Omega-6 | 86155 (99.9%) | 4379 (99.9%) |
| Omega-6-by-Omega-3 | 86152 (99.9%) | 4379 (99.9%) |
| Omega-6-pct | 86155 (99.9%) | 4379 (99.9%) |
| PUFA | 86155 (99.9%) | 4379 (99.9%) |
| PUFA-by-MUFA | 86155 (99.9%) | 4379 (99.9%) |
| PUFA-pct | 86155 (99.9%) | 4379 (99.9%) |
| Phe | 86192 (100.0%) | 4358 (99.4%) |
| Phosphatidylc | 86155 (99.9%) | 4379 (99.9%) |
| Phosphoglyc | 86155 (99.9%) | 4379 (99.9%) |
| Pyruvate | 85994 (99.7%) | 4329 (98.8%) |
| Remnant-C | 86232 (100.0%) | 4383 (100.0%) |
| SFA | 86155 (99.9%) | 4379 (99.9%) |
| SFA-pct | 86155 (99.9%) | 4379 (99.9%) |
| S-HDL-C | 86232 (100.0%) | 4383 (100.0%) |
| S-HDL-CE | 86232 (100.0%) | 4383 (100.0%) |
| S-HDL-CE-pct | 86232 (100.0%) | 4383 (100.0%) |
| S-HDL-C-pct | 86232 (100.0%) | 4383 (100.0%) |
| S-HDL-FC | 86232 (100.0%) | 4383 (100.0%) |
| S-HDL-FC-pct | 86232 (100.0%) | 4383 (100.0%) |
| S-HDL-L | 86232 (100.0%) | 4383 (100.0%) |
| S-HDL-P | 86232 (100.0%) | 4383 (100.0%) |
| S-HDL-PL | 86232 (100.0%) | 4383 (100.0%) |
| S-HDL-PL-pct | 86232 (100.0%) | 4383 (100.0%) |
| S-HDL-TG | 86232 (100.0%) | 4383 (100.0%) |
| S-HDL-TG-pct | 86232 (100.0%) | 4383 (100.0%) |
| S-LDL-C | 86232 (100.0%) | 4383 (100.0%) |
| S-LDL-CE | 86232 (100.0%) | 4383 (100.0%) |
| S-LDL-CE-pct | 86232 (100.0%) | 4383 (100.0%) |
| S-LDL-C-pct | 86232 (100.0%) | 4383 (100.0%) |
| S-LDL-FC | 86232 (100.0%) | 4383 (100.0%) |
| S-LDL-FC-pct | 86232 (100.0%) | 4383 (100.0%) |
| S-LDL-L | 86232 (100.0%) | 4383 (100.0%) |
| S-LDL-P | 86232 (100.0%) | 4383 (100.0%) |
| S-LDL-PL | 86232 (100.0%) | 4383 (100.0%) |
| S-LDL-PL-pct | 86232 (100.0%) | 4383 (100.0%) |
| S-LDL-TG | 86232 (100.0%) | 4383 (100.0%) |
| S-LDL-TG-pct | 86232 (100.0%) | 4383 (100.0%) |
| S-VLDL-C | 86232 (100.0%) | 4383 (100.0%) |
| S-VLDL-CE | 86232 (100.0%) | 4383 (100.0%) |
| S-VLDL-CE-pct | 86232 (100.0%) | 4383 (100.0%) |
| S-VLDL-C-pct | 86232 (100.0%) | 4383 (100.0%) |
| S-VLDL-FC | 86232 (100.0%) | 4383 (100.0%) |
| S-VLDL-FC-pct | 86232 (100.0%) | 4383 (100.0%) |
| S-VLDL-L | 86232 (100.0%) | 4383 (100.0%) |
| S-VLDL-P | 86232 (100.0%) | 4383 (100.0%) |
| S-VLDL-PL | 86232 (100.0%) | 4383 (100.0%) |
| S-VLDL-PL-pct | 86232 (100.0%) | 4383 (100.0%) |
| S-VLDL-TG | 86232 (100.0%) | 4383 (100.0%) |
| S-VLDL-TG-pct | 86232 (100.0%) | 4383 (100.0%) |
| Sphingomyelins | 86154 (99.9%) | 4379 (99.9%) |
| TG-by-PG | 86155 (99.9%) | 4379 (99.9%) |
| Total-BCAA | 86162 (99.9%) | 4374 (99.8%) |
| Total-C | 86232 (100.0%) | 4383 (100.0%) |
| Total-CE | 86232 (100.0%) | 4383 (100.0%) |
| Total-FA | 86155 (99.9%) | 4379 (99.9%) |
| Total-FC | 86232 (100.0%) | 4383 (100.0%) |
| Total-L | 86232 (100.0%) | 4383 (100.0%) |
| Total-P | 86232 (100.0%) | 4383 (100.0%) |
| Total-PL | 86232 (100.0%) | 4383 (100.0%) |
| Total-TG | 86232 (100.0%) | 4383 (100.0%) |
| Tyr | 86131 (99.9%) | 4354 (99.3%) |
| Unsaturation | 86155 (99.9%) | 4379 (99.9%) |
| VLDL-C | 86232 (100.0%) | 4383 (100.0%) |
| VLDL-CE | 86232 (100.0%) | 4383 (100.0%) |
| VLDL-FC | 86232 (100.0%) | 4383 (100.0%) |
| VLDL-L | 86232 (100.0%) | 4383 (100.0%) |
| VLDL-P | 86232 (100.0%) | 4383 (100.0%) |
| VLDL-PL | 86232 (100.0%) | 4383 (100.0%) |
| VLDL-TG | 86232 (100.0%) | 4383 (100.0%) |
| VLDL-size | 86232 (100.0%) | 4383 (100.0%) |
| Val | 86163 (99.9%) | 4373 (99.8%) |
| XL-HDL-C | 86232 (100.0%) | 4383 (100.0%) |
| XL-HDL-CE | 86232 (100.0%) | 4383 (100.0%) |
| XL-HDL-CE-pct | 86232 (100.0%) | 4383 (100.0%) |
| XL-HDL-C-pct | 86232 (100.0%) | 4383 (100.0%) |
| XL-HDL-FC | 86232 (100.0%) | 4383 (100.0%) |
| XL-HDL-FC-pct | 86232 (100.0%) | 4383 (100.0%) |
| XL-HDL-L | 86232 (100.0%) | 4383 (100.0%) |
| XL-HDL-P | 86232 (100.0%) | 4383 (100.0%) |
| XL-HDL-PL | 86232 (100.0%) | 4383 (100.0%) |
| XL-HDL-PL-pct | 86232 (100.0%) | 4383 (100.0%) |
| XL-HDL-TG | 86232 (100.0%) | 4383 (100.0%) |
| XL-HDL-TG-pct | 86232 (100.0%) | 4383 (100.0%) |
| XL-VLDL-C | 86232 (100.0%) | 4383 (100.0%) |
| XL-VLDL-CE | 86232 (100.0%) | 4383 (100.0%) |
| XL-VLDL-CE-pct | 86232 (100.0%) | 4383 (100.0%) |
| XL-VLDL-C-pct | 86232 (100.0%) | 4383 (100.0%) |
| XL-VLDL-FC | 86232 (100.0%) | 4383 (100.0%) |
| XL-VLDL-FC-pct | 86232 (100.0%) | 4383 (100.0%) |
| XL-VLDL-L | 86232 (100.0%) | 4383 (100.0%) |
| XL-VLDL-P | 86232 (100.0%) | 4383 (100.0%) |
| XL-VLDL-PL | 86232 (100.0%) | 4383 (100.0%) |
| XL-VLDL-PL-pct | 86232 (100.0%) | 4383 (100.0%) |
| XL-VLDL-TG | 86232 (100.0%) | 4383 (100.0%) |
| XL-VLDL-TG-pct | 86232 (100.0%) | 4383 (100.0%) |
| XS-VLDL-C | 86232 (100.0%) | 4383 (100.0%) |
| XS-VLDL-CE | 86232 (100.0%) | 4383 (100.0%) |
| XS-VLDL-CE-pct | 86232 (100.0%) | 4383 (100.0%) |
| XS-VLDL-C-pct | 86232 (100.0%) | 4383 (100.0%) |
| XS-VLDL-FC | 86232 (100.0%) | 4383 (100.0%) |
| XS-VLDL-FC-pct | 86232 (100.0%) | 4383 (100.0%) |
| XS-VLDL-L | 86232 (100.0%) | 4383 (100.0%) |
| XS-VLDL-P | 86232 (100.0%) | 4383 (100.0%) |
| XS-VLDL-PL | 86232 (100.0%) | 4383 (100.0%) |
| XS-VLDL-PL-pct | 86232 (100.0%) | 4383 (100.0%) |
| XS-VLDL-TG | 86232 (100.0%) | 4383 (100.0%) |
| XS-VLDL-TG-pct | 86232 (100.0%) | 4383 (100.0%) |
| XXL-VLDL-C | 86232 (100.0%) | 4383 (100.0%) |
| XXL-VLDL-CE | 86232 (100.0%) | 4383 (100.0%) |
| XXL-VLDL-CE-pct | 86232 (100.0%) | 4383 (100.0%) |
| XXL-VLDL-C-pct | 86232 (100.0%) | 4383 (100.0%) |
| XXL-VLDL-FC | 86232 (100.0%) | 4383 (100.0%) |
| XXL-VLDL-FC-pct | 86232 (100.0%) | 4383 (100.0%) |
| XXL-VLDL-L | 86232 (100.0%) | 4383 (100.0%) |
| XXL-VLDL-P | 86232 (100.0%) | 4383 (100.0%) |
| XXL-VLDL-PL | 86232 (100.0%) | 4383 (100.0%) |
| XXL-VLDL-PL-pct | 86232 (100.0%) | 4383 (100.0%) |
| XXL-VLDL-TG | 86232 (100.0%) | 4383 (100.0%) |
| XXL-VLDL-TG-pct | 86232 (100.0%) | 4383 (100.0%) |
| bOHbutyrate | 84559 (98.1%) | 4347 (99.2%) |
| Non-HDL-C | 86232 (100.0%) | 4383 (100.0%) |

**Abbreviations:** Ala, alanine; Apo-A1=apolipoprotein A1; Apo-B=apolipoprotein B; bOHbutyrate, 3-hydroxybutyrate; BCAA= Branched-Chain Amino Acids; C=cholesterol; CE=cholesteryl esters; DHA=docosahexaenoic acid; FA=fatty acids; Gln, glutamine; FC= free cholesterol; Gly, glycine; GlycA, glycoprotein acetyls; HDL=high density lipoproteins; HDL-D=high density lipoprotein particle diameter; His, histidine; IDL=intermediate density lipoproteins; Ile, isoleucine; L=large; LA=linoleic acid; LDL=low density lipoproteins; LDL-D=low density lipoprotein particle diameter; Leu, leucine; LP=lipoprotein; M=medium; MUFA=monounsaturated fatty acids; P=particles; Phe, phenylalanine; PL=Phospholipids; PUFA=polyunsaturated fatty acids; pct= percentage; S=small; SFA=saturated fatty acids; TG=triglycerides; Tyr, tyrosine; Val, valine; VLDL=very low density lipoproteins; VLDL-D=very low density lipoprotein particle diameter; XL=very large; XS=very small; XXL=extremely large.

**Supplemental** **Table S2.** Diagnosis and medication codes for assessment of type 2 diabetes status in primary and secondary healthcare and death registry records and verbal interview

| **Code format** | **Codes** |
| --- | --- |
| ***Diagnosis codes*** | |
| **ICD-10** | E10-E14 |
| ***Medication codes*** | |
| **ATC codes** | A10AB01, A10AB02, A10AB03, A10AB04, A10AB05, A10AB06, A10AB30, A10AC01, A10AC02, A10AC03, A10AC04, A10AC30, A10AD01, A10AD02, A10AD03, A10AD04, A10AD05, A10AD30, A10AE01, A10AE02, A10AE03, A10AE04, A10AE05, A10AE30, A10AF01, A10BA01, A10BA02, A10BA03, A10BB01, A10BB02, A10BB03, A10BB04, A10BB05, A10BB06, A10BB07, A10BB08, A10BB09, A10BB10, A10BB11, A10BB12, A10BB31, A10BC01, A10BD01, A10BD02, A10BD03, A10BD04, A10BD05, A10BD06, A10BD07, A10BD08, A10BD09, A10BD10, A10BD11, A10BF01, A10BF02, A10BF03, A10BG01, A10BG02, A10BG03, A10BH01, A10BH02, A10BH03, A10BH04, A10BH05, A10BJ01, A10BJ02, A10BJ03, A10BJ04, A10BJ05, A10BJ06, A10BK01, A10BK02, A10BK03, A10BX01, A10BX02, A10BX03, A10BX05, A10BX06, A10BX08, A10XA01 |
| **Verbal interview^a^** | 1140857494, 1140857496, 1140857500, 1140857502, 1140857506, 1140857584, 1140857586, 1140857590, 1140868902, 1140868908, 1140874646, 1140874650, 1140874652, 1140874658, 1140874660, 1140874664, 1140874666, 1140874674, 1140874678, 1140874680, 1140874686, 1140874690, 1140874706, 1140874712, 1140874716, 1140874718, 1140874724, 1140874726, 1140874728, 1140874732, 1140874736, 1140874740, 1140874744, 1140874746, 1140882964, 1140883066, 1140884600, 1140910564, 1140910566, 1140910818, 1140921964, 1141152590, 1141153254, 1141153262, 1141156984, 1141157284, 1141168660, 1141168668, 1141169504, 1141171508, 1141171646, 1141171652, 1141173786, 1141173882, 1141177600, 1141177606, 1141189090, 1141189094 |

^a^ Only assessed in UK Biobank. The medication codes for the verbal interview in UKB and details of the corresponding medications can be viewed at: <https://biobank.ndph.ox.ac.uk/ukb/field.cgi?id=20003>

**Abbreviations:** ATC, Anatomical Therapeutic Chemical; ICD-10, the International Statistical Classification of Diseases and Related Health Problems 10th Revision.

**Supplemental** **Table S3.** Hazard ratios per standard deviation increments with 95% confidence intervals and p-values for the associations of the selected metabolites for incident diabetes prediction in the internal validation cohort (30% of UK Biobank, N=25,870) and external validation cohort (ESTHER, N=4,383)

| **Metabolites** | **Internal validation cohort** | | |  | **External validation cohort** | | |
| --- | --- | --- | --- | --- | --- | --- | --- |
|  | **SD** | **HR (95%CI)** | ***P*-value** |  | **SD** | **HR (95%CI)** | ***P*-value** |
| 3-Hydroxybutyrate | 0.063 mmol/L | 0.99 (0.92, 1.05) | 0.671 |  | 0.059 mmol/L | 1.22 (1.13, 1.30) | <0.001 |
| Acetate | 0.035 mmol/L | 1.01 (0.95, 1.06) | 0.798 |  | 0.062 mmol/L | 0.93 (0.82, 1.05) | 0.247 |
| Citrate | 0.013 mmol/L | 1.10 (1.03, 1.17) | 0.003 |  | 0.016 mmol/L | 1.04 (0.95, 1.14) | 0.425 |
| Glutamine | 0.083 mmol/L | 0.93 (0.87, 0.99) | 0.017 |  | 0.106 mmol/L | 0.87 (0.80, 0.95) | 0.002 |
| Glucose | 0.863 mmol/L | 1.49 (1.41, 1.57) | <0.001 |  | 1.478 mmol/L | 1.63 (1.44, 1.85) | <0.001 |
| IDL-CE-pct | 2.369 % | 0.76 (0.72, 0.80) | <0.001 |  | 2.548 % | 0.78 (0.72, 0.84) | <0.001 |
| LA-pct | 3.325 % | 0.76 (0.72, 0.80) | <0.001 |  | 3.574 % | 0.77 (0.71, 0.83) | <0.001 |
| Lactate | 1.072 mmol/L | 1.21 (1.14, 1.29) | <0.001 |  | 3.351 mmol/L | 1.25 (1.13, 1.38) | <0.001 |
| M-LDL-TG-pct | 1.581 % | 1.34 (1.27, 1.40) | <0.001 |  | 2.075 % | 1.38 (1.25, 1.53) | <0.001 |
| Pyruvate | 0.028 mmol/L | 1.15 (1.07, 1.23) | <0.001 |  | 0.068 mmol/L | 1.21 (0.94, 1.56) | 0.139 |
| Tyrosine | 0.014 mmol/L | 1.19 (1.12, 1.26) | <0.001 |  | 0.015 mmol/L | 1.24 (1.14, 1.36) | <0.001 |

**Abbreviations:** CI, confidence interval; IDL-CE-pct, cholesteryl esters to total lipids in IDL percentage; LA-pct, linoleic acid to total fatty acids percentage; M-LDL-TG-pct, triglycerides to total lipids in medium LDL percentage; SD, standard deviation.

### **Supplemental** **Table S4.** ß-coefficients of the variables of the UKB-DRS for 10-year prediction of type 2 diabetes

| **Risk factor (units)** | **ß-coefficient** |
| --- | --- |
| **Clinical CDRS variables** | |
| Female | -0.2878 |
| Prescribed anti-hypertension medication | 0.1046 |
| Prescribed steroids | 0.3950 |
| Age (per years) | 0.1018 |
| Body mass index <25 kg/m^2^ | Ref |
| Body mass index 25-27.49 kg/m^2^ | 0.3632 |
| Body mass index 27.5-29.99 kg/m^2^ | 0.6876 |
| Body mass index ≥ 30 kg/m^2^ | 1.2019 |
| No first degree relative with diabetes | Ref |
| Parent or sibling with diabetes | 0.5114 |
| Parent and sibling with diabetes | 0.7203 |
| None-smoker | Ref |
| Ex-smoker | 0.1297 |
| Current smoker | 0.3409 |
| HbA_1c_ | 0.1014 |
| **Added metabolites** |  |
| 3-Hydroxybutyrate (per 1 SD^a^) | 0.0493 |
| Acetate (per 1 SD^a^) | -0.0214 |
| Citrate (per 1 SD^a^) | -0.0286 |
| Glutamine (per 1 SD^a^) | -0.1055 |
| Glucose (per 1 SD^a^) | 0.3880 |
| IDL-CE-pct (per 1 SD^a^) | -0.1233 |
| LA-pct (per 1 SD^a^) | -0.0691 |
| Lactate (per 1 SD^a^) | 0.1707 |
| M-LDL-TG-pct (per 1 SD^a^) | 0.1300 |
| Pyruvate (per 1 SD^a^) | -0.0454 |
| Tyrosine (per 1 SD^a^) | 0.0458 |

^a^ The standard deviations of the 11 selected metabolites can be found in **Supplemental Table S3**.

**Abbreviations:** CI, confidence interval; CDRS, clinical Cambridge Diabetes Risk Score; IDL-CE-pct, cholesteryl esters to total lipids in IDL percentage; LA-pct, linoleic acid to total fatty acids percentage; M-LDL-TG-pct, triglycerides to total lipids in medium LDL percentage; Ref, reference; SD, standard deviation.

**Supplemental** **Table S5.** Comparison of 11 selected metabolites levels by fasting status in the UK Biobank and ESTHER study

| **Metabolites** | **UK Biobank (N=86,232)** | |  | **ESTHER (N=4,383)** | |
| --- | --- | --- | --- | --- | --- |
|  | **Fasting**  **(N=2,882)** | **Non-fasting**  **(N=83,350)** |  | **Fasting**  **(N=3,940)** | **Non-fasting**  **(N=443)** |
|  | **Mean (95% CI)** | **Mean (95% CI)** |  | **Mean (95% CI)** | **Mean (95% CI)** |
| 3-Hydroxybutyrate, mmol/L | 0.081 (0.077, 0.084) | 0.062 (0.062, 0.062) |  | 0.070 (0.069, 0.072) | 0.063 (0.058, 0.068) |
| Acetate, mmol/L | 0.023 (0.021, 0.025) | 0.018 (0.018, 0.018) |  | 0.040 (0.038, 0.041) | 0.046 (0.034, 0.057) |
| Citrate, mmol/L | 0.067 (0.067, 0.068) | 0.066 (0.066, 0.066) |  | 0.059 (0.059, 0.060) | 0.062 (0.060, 0.064) |
| Glutamine, mmol/L | 0.554 (0.551, 0.557) | 0.558 (0.558, 0.559) |  | 0.549 (0.545, 0.552) | 0.554 (0.544, 0.565) |
| Glucose, mmol/L | 3.66 (3.63, 3.68) | 3.64 (3.64, 3.65) |  | 3.83 (3.79, 3.88) | 3.94 (3.77, 4.12) |
| IDL-CE-pct, % | 50.2 (50.1, 50.3) | 50.1 (50.1, 50.1) |  | 50.5 (50.4, 50.6) | 50.1 (49.8, 50.4) |
| LA-pct, % | 28.8 (28.7, 28.9) | 28.8 (28.8, 28.8) |  | 30.2 (30.1, 30.3) | 29.6 (29.2, 29.9) |
| Lactate, mmol/L | 3.93 (3.90, 3.98) | 3.84 (3.84, 3.86) |  | 5.46 (5.36, 5.57) | 6.10 (5.76, 6.43) |
| M-LDL-TG-pct, % | 5.19 (5.13, 5.26) | 5.42 (5.41, 5.43) |  | 5.46 (5.40, 5.53) | 6.07 (5.86, 6.28) |
| Pyruvate, mmol/L | 0.078 (0.077, 0.079) | 0.079 (0.079, 0.079) |  | 0.095 (0.093, 0.097) | 0.102 (0.095, 0.109) |
| Tyrosine, mmol/L | 0.058 (0.058, 0.059) | 0.064 (0.064, 0.064) |  | 0.069 (0.069, 0.070) | 0.076 (0.074, 0.078) |

**Abbreviations:** CI, confidence interval; IDL-CE-pct, cholesteryl esters to total lipids in IDL percentage; LA-pct, linoleic acid to total fatty acids percentage; M-LDL-TG-pct, triglycerides to total lipids in medium LDL percentage; SD, standard deviation.

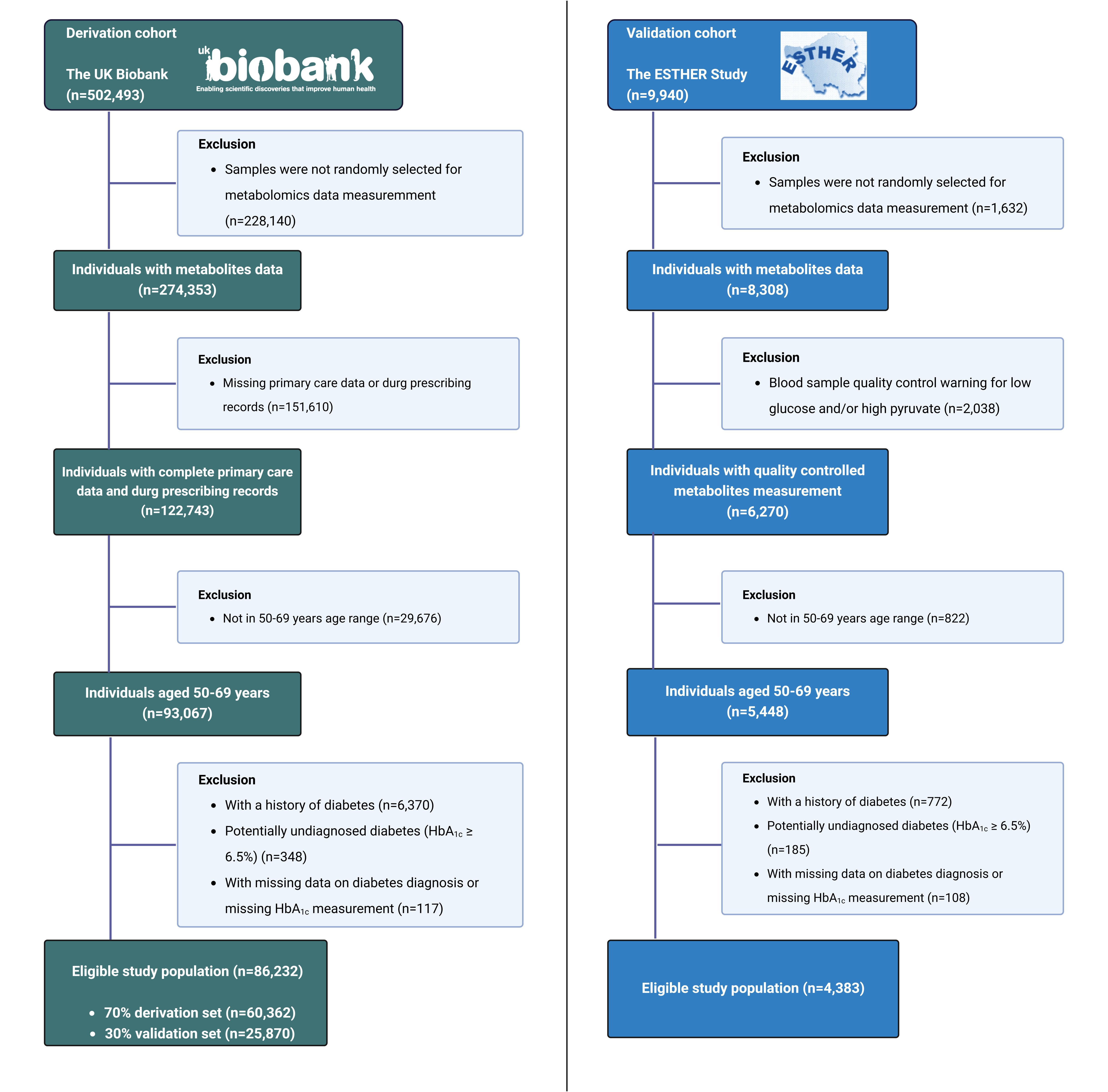

### **Supplemental** **Figure S1.** Flow-charts for participant inclusion and exclusion

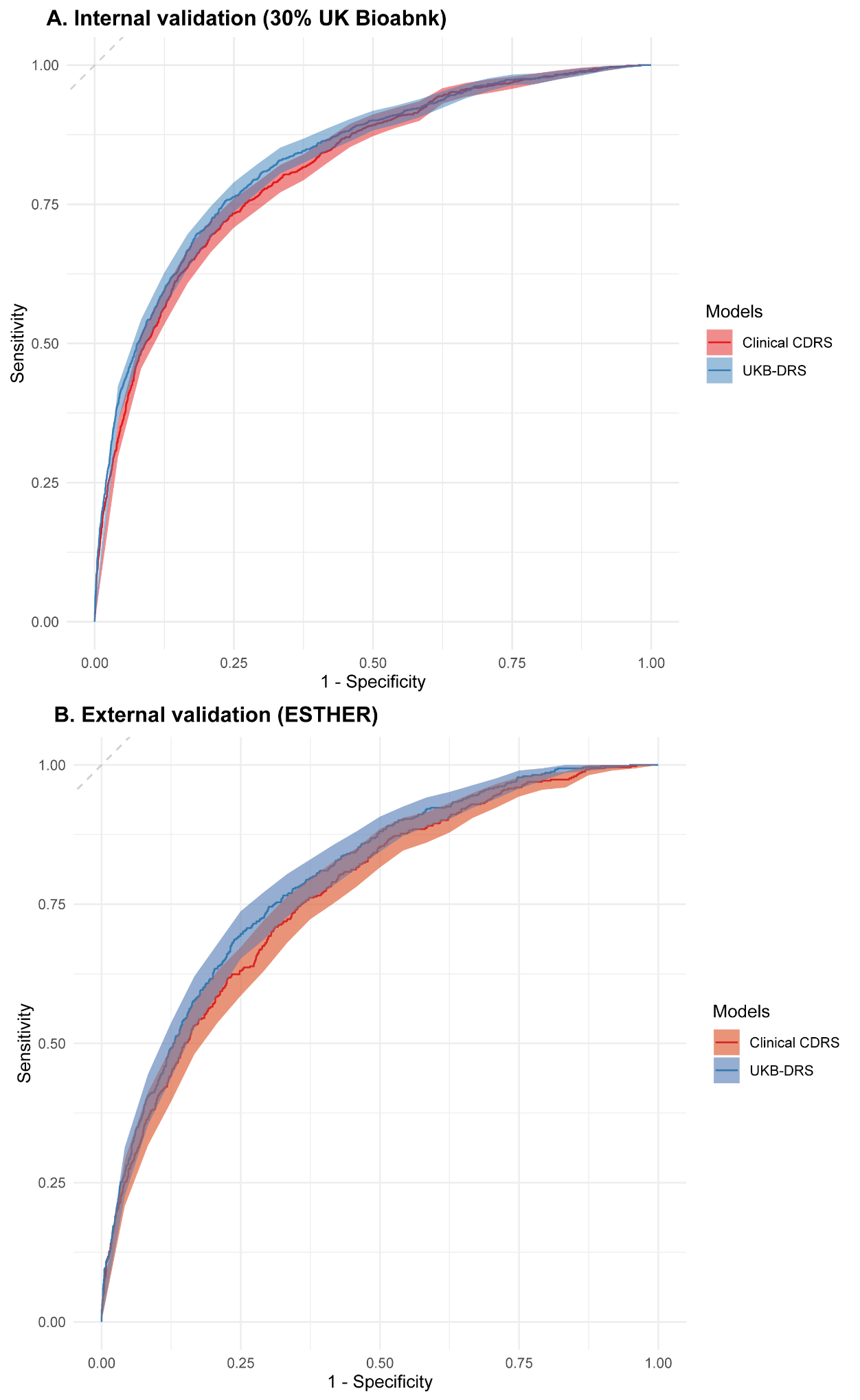

### **Supplemental Figure S2.** ROC curves of the clinical CDRS and the UKB-DRS for 10-year type 2 diabetes risk prediction in the internal validation (30% of UK Biobank, N=25,870) and external validation (ESTHER, N=4,383)

Metabolites that were included in the UKB-DRS were 3-hydroxybutyrate, acetate, glutamine, glucose, IDL-CE-pct, LA-pct, lactate, M-LDL-TG-pct, pyruvate and tyrosine (see **Supplemental Table S3** for abbreviations).

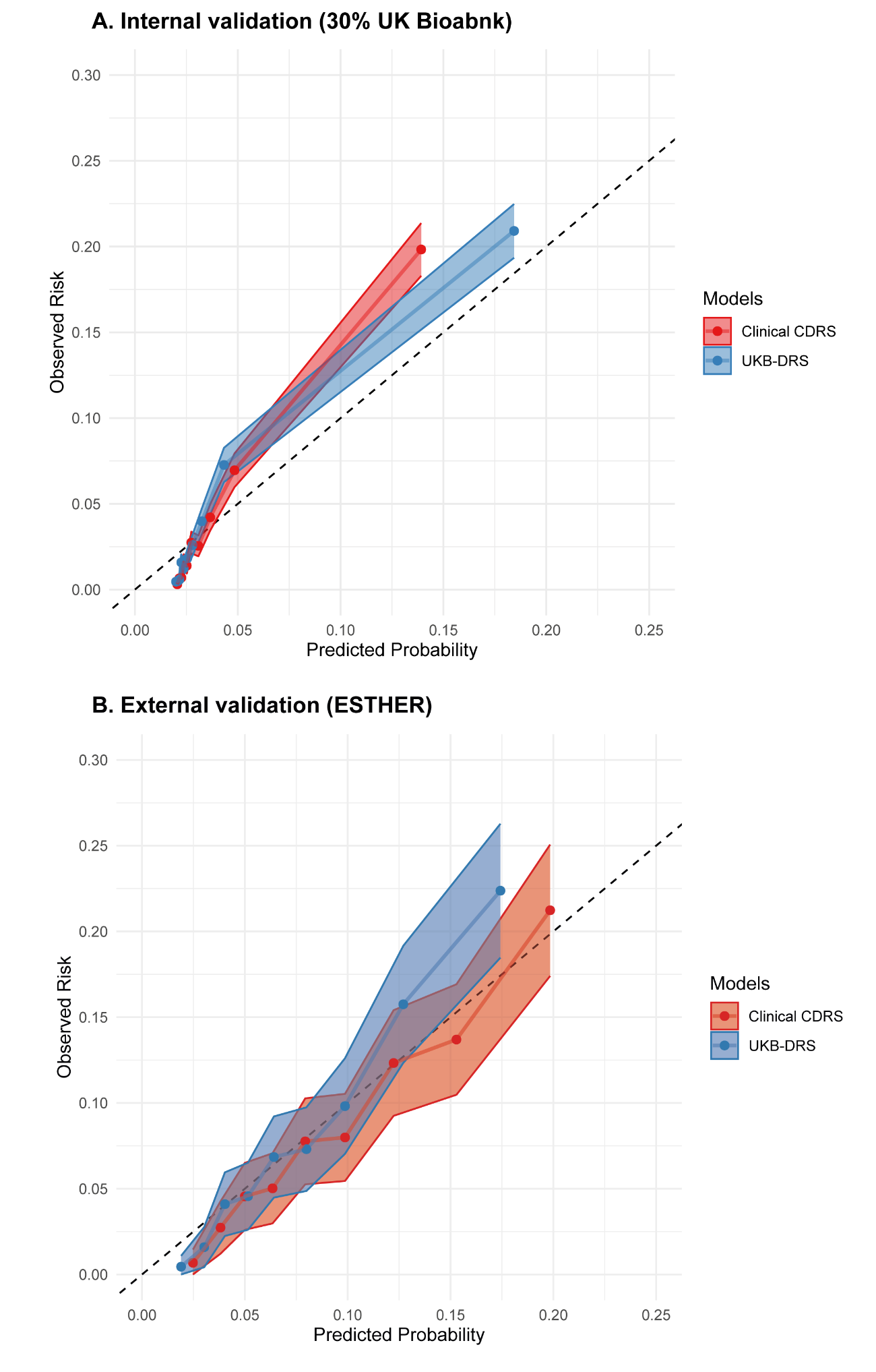

### **Supplemental Figure S3.** Calibration curves of the clinical CDRS and the UKB-DRS for 10-year type 2 diabetes risk prediction in the internal validation (30% of UK Biobank, N=25,870) and external validation (ESTHER, N=4,383)

Metabolites that were included in the UKB-DRS were 3-hydroxybutyrate, acetate, glutamine, glucose, IDL-CE-pct, LA-pct, lactate, M-LDL-TG-pct, pyruvate and tyrosine (see **Supplemental Table S3** for abbreviations).
